# Muscle Ultrasound Image Analysis with a Semi-Automated Software (MuscleExpert) Yields Results Comparable to ImageJ: Muscle Thickness, Muscle Area, and Muscle Quality

**DOI:** 10.1101/2025.09.01.25334864

**Authors:** Marcel Bahia Lanza

**Author notes:** **Corresponding Author:** Marcel Bahia Lanza Ph.D.; Department of Physical Therapy and Rehabilitation Science, School of Medicine, University of Maryland Baltimore, 100 Penn Street Baltimore, MD 21201- 1082, United States, United States.

## Abstract

**Purpose:** The aim of this study is to present and validate the semi-automated MuscleExpert software against the widely used ImageJ by comparing muscle thickness (MT), muscle area (MA), and muscle quality (MQ-measured as grayscale).

**Methods:** Ten volunteers participated in this study. Ultrasound images of the tibialis anterior and medial gastrocnemius muscles were acquired using 2D B-mode ultrasonography and analyzed on both software. Equivalence of means was evaluated via the two‐one‐sided tests (TOST) procedure, the reliability between means was quantified by an intraclass correlation coefficient (ICC), the precision was expressed as the coefficient of variation as well as standard error of measurement (SEM), and the agreement across the measurement was assessed using Bland–Altman plots.

**Results:** The TOST for MT, MA, and MQ was within their respective equivalence margins. The ICC was excellent for all measures (MT: ICC=0.999; MA: ICC=0.999; MQ: ICC=1.000). Precision was similarly high, with CVs of 0.49% for MT, 0.54% for MA, and 0.33% for MQ, demonstrating minimal variability between the two methods. SEM values were low across all outcomes, corresponding to 0.49 % for MT, 0.53% for MA, and 0.23% for MQ. Finally, the Bland–Altman analysis demonstrated minimal systematic differences and no proportional bias between ImageJ and MuscleExpert for all three metrics.

**Conclusion:** The present findings indicate that MuscleExpert and ImageJ produce nearly identical results for MT, MA, and MQ. These findings support the integration of MuscleExpert into clinical and research workflows, offering a more efficient solution for muscle assessment without compromising measurement integrity.

## Introduction

Two-dimensional (2D) ultrasound (US) imaging is widely used for clinical and research purposes across health sciences field^1^. Clinicians often record images of organs to assess structural size and tissue quality^2,3^, while researchers and physical trainers use B-mode ultrasound to capture images of specific muscles and quantify both muscle size^4^ and muscle quality (e.g., grayscale)^5^. The accuracy and consistency of ultrasound-derived measurements can vary depending on the analysis software and user workflow^6^. To ensure reliable and reproducible results, as well as decreased time for data analysis, there is a clear need for software specifically designed for ultrasound image analysis.

Most image‐analysis software—such as Horos, OsiriX, and ImageJ—were not specifically designed for ultrasound‐based muscle morphology measurements. Consequently, they contain numerous general‐purpose features that are irrelevant to this application and force users to navigate through multiple menus and dialogs. In many cases, the overwheliming amount of tools can prolong data analysis and may lead users to select the wrong functions, resulting in measurement errors^7^. Thus, this maybe the reason for researchers to develop their own custom image-analysis software and/or scripts^8,9^. However, these tools are rarely shared with the wider community, and there’s no assurance that the measurements they produce will agree with those generated by established, widely used programs. Consequently, it’s possible that measurements derived from such tools differ meaningfully from those obtained using standard, well-documented platforms—introducing potential bias or error into published findings.

Muscle size and quality are types of measurements that are constantly performed by different research groups using ultrasound^4,10–14^. The image analysis to extract the values from these measurements, can be time consuming. For reference, a typical analysis on ImageJ software to extract both measurements (muscle size and quality) proceeds through the following steps: 1) loading the raw ultrasound image; 2) opening the calibration tool; 3) drawing a reference line over a region of known length (i.e., the probe’s field of view); 4) entering the corresponding distance to set the image scale; 5) opening brightness/contrast settings (not always required); 6) adjusting brightness and/or contrast (not always required); 7) applying image filters to enhance visualization of superficial and deep aponeuroses (not always required); 8) selecting the muscle‐thickness measurement tool; 9) tracing a line between the superficial and deep aponeuroses of the muscle; 10) recording the value; 11) selecting the polygon tool to draw the region of interest (ROI) for echo‐intensity analysis; 12) trace the ROI; 13) extracting the mean grayscale value; and 14) saving or exporting the measurement data. Switching between these tools for each task not only prolongs the process but also increases the risk of inconsistency and user error.

Recently, the MuscleExpert software was released freely on Zenode^15^. This semi-automatic python based program was designed to reduce analysis time by minimizing manual steps and automatically selecting the appropriate tools based on established muscle-morphology protocols across different research groups^10,16–19^. Unlike ImageJ, MuscleExpert streamlines analysis into four steps: 1) loading the raw US image; 2) cropping the image of the muscle; 3) drawing the ROI. By collapsing more than a dozen manual operations into just these three actions, MuscleExpert slashes analysis time by at least 80% and eliminates the risk of tool-selection errors, ensuring faster, and potentially more consistent measurements across users. However, the reproducibility and reliability of MuscleExpert have not been demonstrated, nor has its agreement with other well-known software.

Therefore, the aim of this study is to validate the semi-automated MuscleExpert software against the widely used ImageJ software by comparing muscle thickness (MT), muscle area (MA), and muscle quality (MQ-measured as grayscale) metrics to assess agreement and reliability between the two approaches. It was hypothesized that the MuscleExpert software would demonstrate excellent agreement and reliability with ImageJ, thereby supporting its adoption as a tool for ultrasound image analysis.

## Methods

### Subjects

This study was approved by the University’s Institutional Review Board. A convenience sample of 10 participants (18 to 85 years old; age: 40.1 ± 24.3 years; height: 1.65 ± 0.8 m; body mass: 67.3 ± 16.4 kg) provided written informed consent before their participation. All participants had no history of neurological or muscular disorder, nor serious injuries or surgery of the lower extremity volunteered to participate in the study.

### Measurements and Procedures

Participants remained seated (knees at 90º of flexion) and rested for 10 min before ultrasound measurements. Ultrasound recordings and analysis were performed for tibialis anterior (TA) and medial gastrocnemius (MG) muscles. The anterior and posterior region of the dominant leg were marked to identify the reference points for the ultrasound image acquisition. The dominant limb was defined by the question: which foot would you kick a ball?^16^. Skin marks were made at 1/3 on the line between the tip of the fibula and the tip of the medial malleolus for the TA muscle, while placed on the most prominent bulge of the muscle for MG. Two ultrasound images per site were obtained using B-mode ultrasonography (Aixplorer, Supersonic Imagine, Aix-en-Provence, France) with a SuperLinear 5-18 MHz frequency probe, gain of 62 dB, a depth of 6.0 cm. An example of US images can be seen in Figure 1.

**Figure 1.**
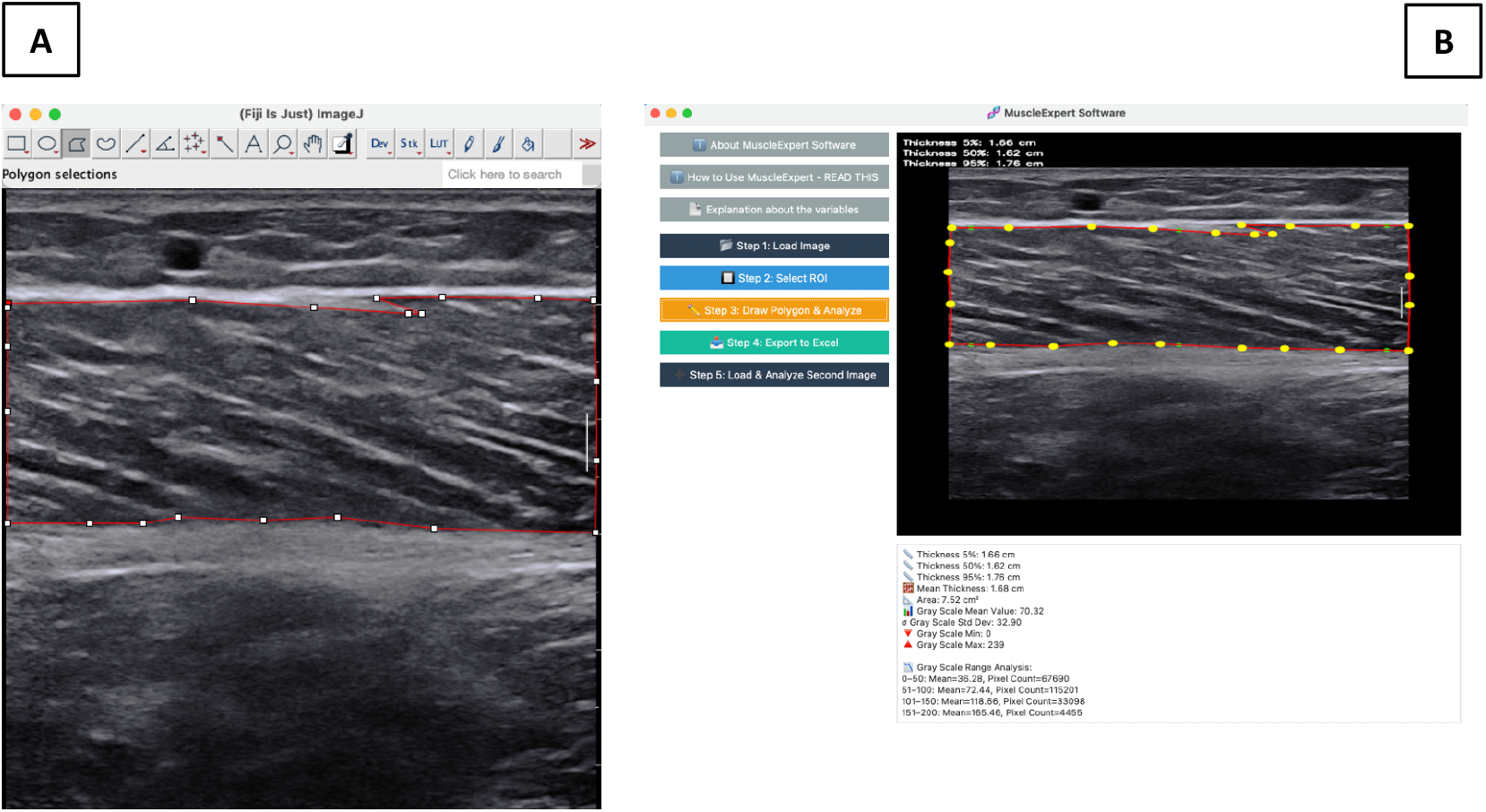
Example of a muscle area measurement from the medial gastrocnemius from one participant on (A) ImageJ software and (B) MuscleExpert software.

### Data analysis

For each muscle, twenty images were generated, yielding a total of forty images for analysis. Image analysis was performed using two software programs: ImageJ software, version 1.53a (National Institutes of Health, USA) and MuscleExpert software, version 1.0. The image analysis comprised two stages. In the first stage, each image was analyzed twice in each program (ImageJ and MuscleExpert), and the mean of those two readings was calculated for each program. In the second stage of analysis, the same procedure was repeated. The grand mean—computed as the average of the two stages—served as the final measurement for all subsequent analyses. Repeating measurements within and between sessions and then averaging them has been shown to reduce random error and to allow formal assessment of intra‐rater consistency using intraclass correlation coefficients^20,21^. Moreover, this design enables the detection of any systematic bias (e.g., learning or fatigue effects) between stages, yielding more precise and trustworthy estimates.

Ultrasound images were exported from the imaging device in JPEG format and imported into both software’s. In ImageJ, MT was measured as the straight‐line distance between the superficial and deep aponeuroses on each image, while muscle quality (mean grayscale value) was determined by outlining the ROI in the TA and MG muscles with the polygon tool^7^ and recording the average grayscale value within that area. In MuscleExpert, the polygon tool was used to delineate muscle ROI, but both MT and mean grayscale were calculated automatically from the drawn region. On both software’s, MT was measured at 50% of the image size as previously^4^.

### Statistical analysis

Statistical analyses were conducted in Python using the Visual Studio Code IDE (Microsoft Corporation, version 1.90.2), and an alpha level of 0.05 was applied to determine statistical significance. Data from the two muscles were pooled because our goal was to compare software performance rather than muscle-specific differences. All variables were assessed for normality using the Shapiro–Wilk test. Equivalence of means was then evaluated via the two‐one‐sided tests (TOST) procedure^22^ with a priori equivalence margins (Δ) of ± 0.25 cm for thickness, ± 0.65 cm^2^ for area, and ± 9 arbitrary units (au.). Two one‐sided paired t‐tests were performed and equivalence concluded if both p‐values were < 0.05 and the 90 % confidence interval for the mean difference lay entirely within [–Δ, +Δ] ^22^. Margins for MT^4,23–26^ and MQ (as grayscale)^24,26–28^ in both muscles were calculated from prior studies. The most conservative margin was then selected—that is, the one corresponding to the muscle with the smallest observed range (which was no more than 8%)—and present it above. Because measuring MA on longitudinal US images is not standard practice—and here was performed to assess agreement between software programs—we imposed an 8% conservative margin based on our ImageJ data. Absolute agreement was quantified by an intraclass correlation coefficient (ICC_(3,1)_), interpreted as weak (<0.4), moderate (0.4-0.59), good (0.6-0.74) and/or excellent (0.75-1.0)^29^. Precision was expressed as the coefficient of variation (CV = [within‐subject SD /overall mean] × 100 %). The standard error of measurement (SEM) was computed using the classical test theory formula: 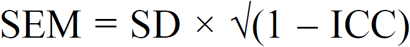, where SD represents the pooled standard deviation of the two measurement methods and ICC refers to the absolute agreement intraclass correlation coefficient (ICC_(3,1)_). SEM was further expressed as a percentage of the overall mean to provide a scale-free index of absolute error. Agreement across the measurement range was further explored using Bland–Altman plots, reporting mean bias and 95 % limits of agreement (mean difference ± 1.96 × SD).

## Results

The TOST for MT yielded a mean difference of 0.005 cm (90 % CI: -0.0004 to -0.010cm; p_lower_ < 0.0001, p_upper_ < 0.0001), which lies entirely within the ±0.25 cm margin, Figure 2A. MA differences averaged 0.031 cm^2^ (90 % CI: -0.007 to -0.054 cm^2^; p_lower_ < 0.0001, p_upper_ < 0.0001; Figure 2B) and MQ differences averaged 1.27 au. (90 % CI: -1.143 to -1.397 au.; p_lower_ < 0.0001, p_upper_ < 0.0001; Figure 2C), both comfortably within their respective equivalence margins. Absolute agreement was excellent for all measures (MT: ICC = 0.999; MA: ICC = 0.999; MQ: ICC = 1.000), Table 1. Precision was similarly high, with CVs of 0.49% for MT, 0.54% for MA, and 0.33% for MQ, demonstrating minimal variability between the two methods. SEM values were low across all outcomes, corresponding to 0.008 cm (0.49 %) for MT, 0.04 cm^2^ (0.53 %) for MA, and 0.36 au. (0.23 %) for MQ, Table 1. Bland–Altman analysis demonstrated minimal systematic differences and no proportional bias between ImageJ and MuscleExpert for all three metrics. For MT bias was 0.0052 cm and 95% limits of –0.0296 to 0.0401 cm (slope = 0.0084, p = 0.2673), Figure 3A. For MA the bias was 0.0309 cm^2^ (95% limits –0.1396 to 0.2014 cm^2^) and a non‐significant slope of 0.0154 (p = 0.0610), Figure 3B. Lastly, MQ bias was 1.2701 au., with limits of 0.3379 to 2.2023 au. and slope –0.0037 (p = 0.4654), Figure 3C.

**Table 1.** Intraclass correlation coefficients (ICC), coefficients of variation (CV), and standard error of measurement (SEM) for MuscleExpert versus ImageJ software.

| Variable | ICC (95% CI) | CV (%) | SEM (95% CI) | SEM (%) |
| --- | --- | --- | --- | --- |
| Muscle Thickness | 0.999 (0.998–1.000) | 0.49 | 0.008 (0.0072 - 0.0103) cm | 0.49% |
| Muscle Area | 0.999 (0.999–1.000) | 0.54 | 0.04 (0.03 - 0.05) cm<sup>2</sup> | 0.53% |
| Muscle Quality | 1.000 (0.999–1.000) | 0.33 | 0.23 (0.20 - 0.26) au. | 0.23% |

**Figure 2.**
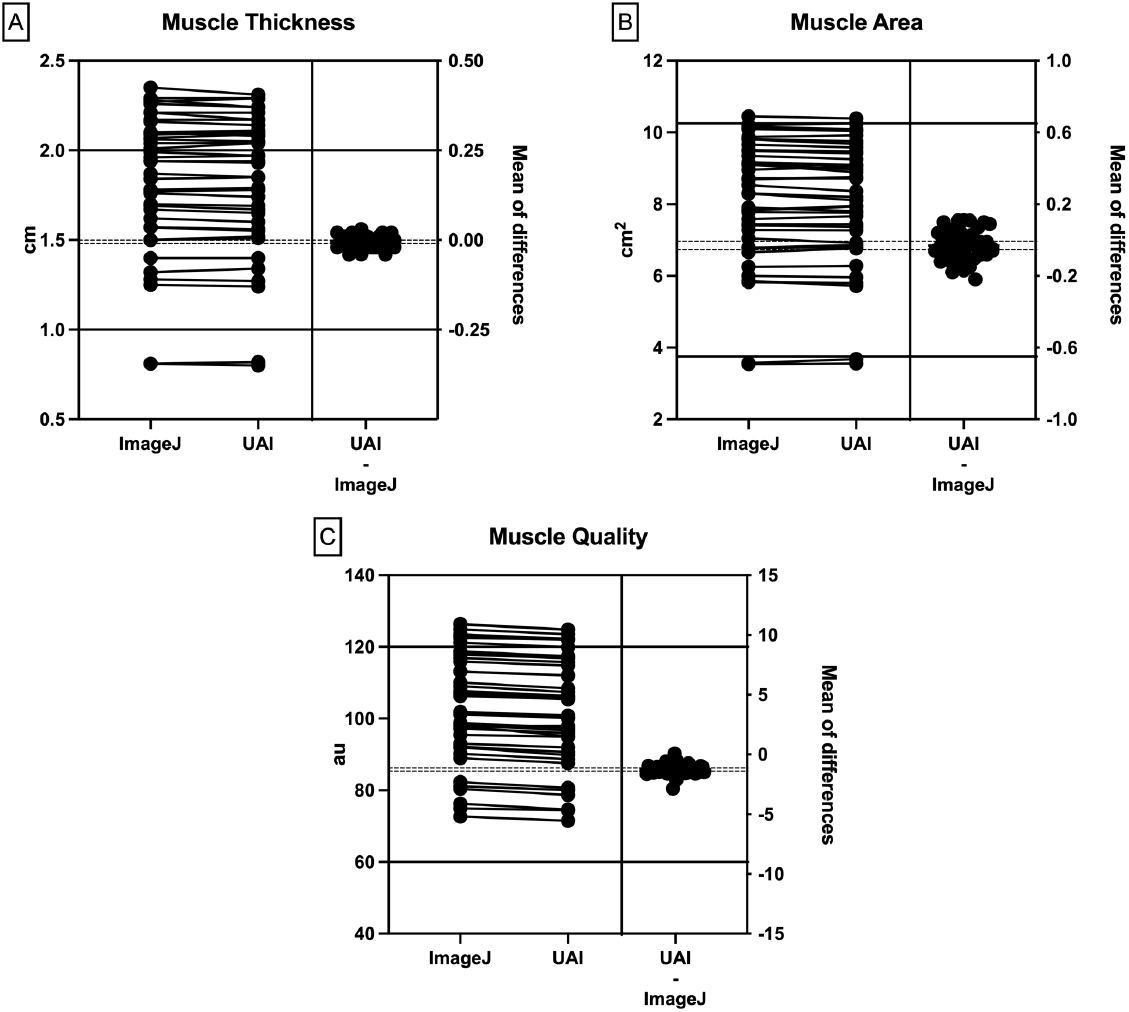
This panel shows mean values of each individual measure on ImageJ and MuscleExpert software’s and the mean‐difference results for (A) muscle thickness, (B) muscle area, and (C) muscle quality. Solid black lines denote the pre‐specified lower (–Δ) and upper (+Δ) equivalence margins used in the two‐one‐sided tests (TOST). Dashed lines indicate the lower and upper bounds of the 90% confidence interval around the observed mean difference. Equivalence is achieved when the entire dashed interval lies within the solid black margins.

**Figure 3.**
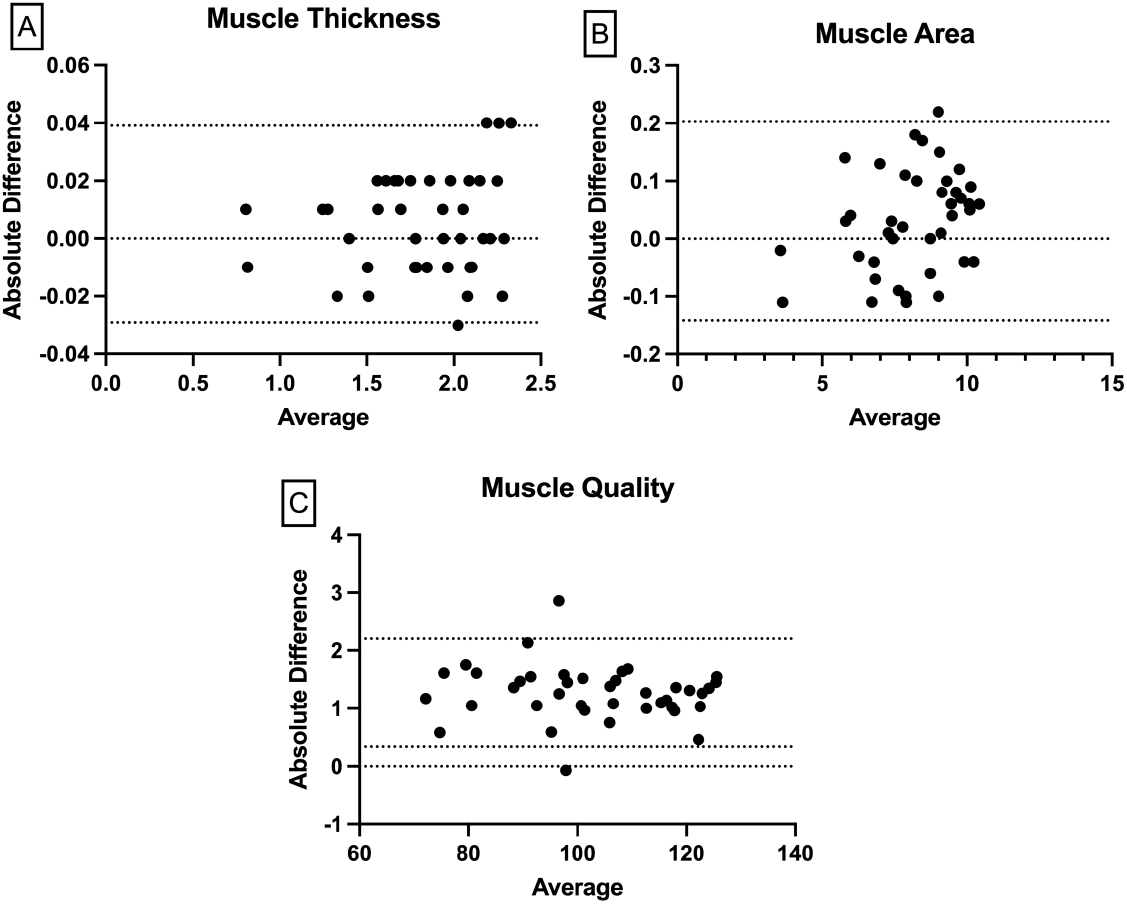
Bland-Altman plots for the inter-rater assessment of muscle thickness (A), muscle area (B), and muscle quality (C), with limits of agreement (dotted line), from -1.96 SD (standard deviation) to +1.96 SD.

## Discussion

Since custom image‐analysis pipelines are often neither published nor benchmarked against established standards, there’s no guarantee their results align with those produced by validated, widely adopted software. Thus, the first aim of this study was to validate the semi-automated MuscleExpert software against ImageJ by comparing MT, MA, and MQ measurements for agreement and reliability. The present results support our hypothesis that the MuscleExpert software demonstrates excellent agreement and reliability with ImageJ. In summary, the MuscleExpert software offers a reliable, efficient alternative to ImageJ for quantitative ultrasound assessment of muscle morphology.

The test of equivalence (TOST) confirmed that the differences in MT, MA, and MQ between the two methods were not only statistically significant but also well within predefined equivalence margins. Importantly, it was adopted highly conservative margins for all variables. This suggests that MuscleExpert can be considered a valid alternative to ImageJ for these measurements. In addition to that, the exceptionally high intraclass correlation coefficients (ICCs ≥ 0.999) and low coefficients of variation (CVs < 1%) further support the precision and consistency of MuscleExpert, indicating minimal variability and excellent reproducibility across all three muscle morphology measurements. SEMs were also consistently low across all outcomes, with values below 0.05% of the pooled mean, further demonstrating the high accuracy of MuscleExpert compared to ImageJ. These minimal SEM values reinforce the notion that the software introduces negligible error. Bland–Altman analyses reinforced these findings by showing minimal systematic bias and no evidence of proportional bias between the two methods. The narrow limits of agreement and non-significant regression slopes across all metrics suggest that MuscleExpert performs comparably to ImageJ across the full range of values. Notably, the MQ metric, often more susceptible to variability due to grayscale interpretation^5^, showed the highest precision (CV = 0.33%) and perfect agreement (ICC = 1.000), highlighting the robustness of MuscleExpert’s grayscale analysis. These findings collectively validate the MuscleExpert software as a reliable and efficient tool for muscle assessment, offering potential advantages in clinical and research settings where automation and scalability are increasingly important.

Each platform presents both advantages and limitations. MuscleExpert is specifically engineered for ultrasound imaging in health‐science disciplines (e.g., exercise physiology, physical therapy), offering a minimalist interface that embeds essential algorithms to eliminate tool‐selection errors and minimize configuration time. In contrast, ImageJ is an open‐source, extensible environment with comprehensive filters, plugins, scripting support (Java, Python), batch‐processing pipelines and detailed metadata extraction. However, Its extensive feature set requires a long period to adatp. Consequently, MuscleExpert is ideal for rapid, error‐resistant standard analyses, whereas ImageJ better serves advanced users requiring deep customization and automation.

In conclusion, the findings indicate that MuscleExpert and ImageJ produce nearly identical results for MT, MA, and MQ. The equivalence testing, high ICC, low Cvs ans SEM values, and Bland–Altman analyses collectively demonstrate that MuscleExpert provides measurements that are statistically and practically indistinguishable from those obtained with ImageJ. These findings support the integration of MuscleExpert into clinical and research workflows, offering a more efficient solution for muscle assessment without compromising measurement integrity.

## Data Availability

Data is available upon reasonable request.

